# Effects of commonly used antibiotics on children’s developing gut microbiomes and resistomes in peri-urban Lima, Peru

**DOI:** 10.1101/2024.12.13.24317790

**Authors:** Neha Sehgal, Monica J. Pajuelo, Robert H. Gilman, Amy J. Pickering, Ashlee M. Earl, Colin J. Worby, Maya L. Nadimpalli

## Abstract

**Background:** The effects of antibiotic use on children’s gut microbiomes and resistomes are not well characterized in middle-income countries, where pediatric antibiotic consumption is exceptionally common. We characterized the effects of antibiotics commonly used by Peruvian children (i.e., amoxicillin, azithromycin, cefalexin, sulfa-trimethoprim) on gut diversity, genera, and antibiotic resistance gene (ARG) abundance from 3-16 months.

**Methods:** This study included 54 children from a prospective cohort of enteric infections in peri-urban Lima, 2016-2019. Stool collected at 3, 6, 7, 9, 12, and 16 months underwent DNA extraction and short-read metagenomic sequencing. We profiled the taxonomy of stool metagenomes and assessed ARG abundance by aligning reads to the ResFinder database. We used daily surveillance data (40,662 observations) to tabulate the number of antibiotic courses consumed in the 30 days prior to stool sampling. Using linear mixed models, the association of recent antibiotic use with species richness, diversity, gut genera, and ARG abundance over time was examined.

**Results:** Most children were vaginally delivered (73%), received breastmilk almost daily over the study period, and belonged to socioeconomically diverse households. Amoxicillin, azithromycin, cefalexin, and sulfa-trimethoprim did not impact gut diversity or genera abundance. Azithromycin use significantly impacted ARGs from the macrolide, aminoglycoside, and folate pathway antagonist classes. Amoxicillin use significantly increased total ARGs. Antibiotics’ effects on ARGs appeared to be independent of gut microbiome changes.

**Conclusion:** Common antibiotics like amoxicillin and azithromycin may be key drivers of the gut resistome but not the microbiome during early childhood in this setting with frequent breastfeeding.

## Introduction

Antibiotic-resistant infections disproportionally occur in low– and middle-income countries (LMICs).^1^ Antibiotic-resistant pediatric infections are especially concerning^1–3^ because alternate treatments may be unavailable or expensive, and these infections have a higher mortality risk.^1,4–6^

Studies in high-income settings have demonstrated that antibiotic administration during pregnancy or early childhood may increase the load of antibiotic resistance genes (ARGs) harbored by gut bacteria,^7–12^ collectively known as the “resistome”, and collaterally, also alter the development of the gut microbiome^11–17^ at an early, dynamic, and sensitive stage of life. Children can be exposed to antibiotics or their residues prenatally via cord blood, and postnatally via breastmilk and direct consumption.^18,10,19^ Children in LMICs frequently consume antibiotics^20^, likely due to higher pathogen exposures, indiscriminate use and poor control of antibiotic usage, but impacts on their gut microbiomes and resistomes are not well-characterized.^21^ Because the microbiome does not stabilize to an “adult-like” state until 2-3 years of age, these frequent perturbations may have acute impacts.

Using data from a prospective study of enteric infections among Peruvian children 0-2 years of age, we examined how recent exposures to commonly used antibiotics altered children’s gut microbiomes and resistomes over the first 16 months of life. We hypothesized that increased antibiotic use would enrich the resistome but decrease the load of gut genera that are often sensitive to antibiotics, such as *Bifidobacterium*.

## Methods

### Study Population

The parent cohort study (NIH R01AI108695-01A1) enrolled 345 children living in Villa El Salvador, Lima between February 2016-May 2019. Weekly stool samples were collected and feeding practices and medication use were surveyed daily. Recruitment methods and enrollment criteria are detailed elsewhere.^22^

As previously described, ^22,23^ a subset of 112 children were screened for fecal carriage of extended spectrum beta lactamase-producing producing Enterobacterales (ESBL-E) from 1-16 months of age. Here, we included 54 children: all 12 children with *rare* ESBL-E gut colonization and a random subset of 42 with *frequent* ESBL-E gut colonization during this period.^23^ Detailed sociodemographic characteristics are available elsewhere.^23^ Briefly, approximately half were female (54%), most were vaginally delivered (72%), and 63% were born to high school-educated mothers.

### Ethical Approval

Infants’ caretakers provided written informed consent for participation in the parent cohort and the use of collected specimens for subsequent research. The Institutional Review Boards (IRBs) of the Universidad Peruana Cayetano Heredia (UPCH), Johns Hopkins University and Asociación Benefica PRISMA approved the parent study. Analyses for this sub-study were approved by the IRBs of UPCH (no. 201592), PRISMA, and Tufts University.

### Exposure Definitions

At each daily survey visit, fieldworkers asked caretakers if the child had consumed any antibiotic or medication in the past 24 hours. If yes, caretakers were asked to provide the packaging so that the fieldworker could confirm the medication type. We defined the start of a new antibiotic course as any timepoint when caretakers first reported antibiotic use following two days of no exposure, and its end when the child did not consume antibiotics for two consecutive days after. We tabulated the number of antibiotic courses in the 30 days prior to a stool sample. For this sub-study, we considered the effects of “commonly used” antibiotics, *i.e.*, those used at least once by ≥10% of 345 children in the parent cohort. These included amoxicillin (14.6 courses/100 child-months), sulfa-trimethoprim (9.4 courses/100 child-months), azithromycin (4.2 courses/100 child-months), and cefalexin (4.1 courses/100 child-months).

Covariate data were obtained from enrollment surveys (*e.g.*, child sex), annual baseline surveys (*e.g.*, maternal education, household poultry ownership), or daily surveys (*e.g.*, recent diarrhea, feeding practices, child age, and time between defecation and diaper retrieval). Child delivery mode was determined by field workers after study completion.

### Metagenome sequencing and resistome profiling

Detailed methods for metagenomic sequencing, quality control, and taxonomic profiling are provided elsewhere.^23^ Briefly, total DNA was extracted from 0.25g of frozen stool collected at 3, 6, 7, 9, 12 and 16 months at UPCH, then shipped on dry ice to the Broad Institute for short-read, paired-end 150bp sequencing using the Illumina Novaseq 6000 System with SP4 flow-cells. Data analyses were performed on the Tufts’ HPC Research Cluster. Sequencing adaptors and low-quality reads were removed using bbmap. MetaPhlAn3 (db v31) was used for taxonomic assignment. The ‘vegan’ package was used to determine species richness and Shannon diversity.^24^

ARGs were identified by mapping short reads to the Resfinder database (v. 3.1.1) using the KMA tool.^25^ Matches with >90% coverage and >95% identity were considered true hits. We identified the number of genome equivalents in each sample using Microbe Census.^26^ To normalize for the number of bacterial genomes in each sample, ARG abundance was calculated as fragments per kilobase per million mapped reads (FPKM) divided by the total number of genome equivalents in that sample.^27^ FPKM for each detected ARG, each ARG class, and overall was determined per sample. FPKMs were log10 transformed for analysis.

### Statistical Analysis

Antibiotic use and diarrheal episodes in the 30 days before a stool sample (hereafter referred to as “recent antibiotic use” or “recent diarrhea”) were summarized using frequencies. Means and standard deviations (SDs) were used to describe continuous measures by child age. T-tests or ANOVAs were conducted to determine differences in means based on child sex, maternal education, and delivery mode.

Samples with a mean number of reads ±2SD were excluded (n=11). To avoid spurious correlations, we only included gut genera, ARG classes, and ARGs detected in >10% of the repeated samples. Nitroimidazole and 146/219 individual ARGs were excluded. Further, because allelic variants of β-lactamase genes can differ by one single nucleotide polymorphism, we considered these genes as groups (e.g., *bla*_CTX_, *bla*_OXA_, *bla*_TEM_) rather than as individual variants in our analyses.

First, we used unadjusted and adjusted linear mixed models to investigate the effect of recent antibiotic use on ARG abundance, including overall, by class, and by individual ARGs. Mixed models were used to account for our inclusion of repeated observations from the same children and estimate a population-level effect that measures the average effect of recent antibiotic use on the outcome while recognizing that each child might have a different baseline and response to antibiotic use over the first 16 months of life (that is, a random effect). Covariates were selected *a priori* based on our direct acyclic graph (**Figure S1**). All adjusted models included recent diarrhea (repeated measurement), child sex (fixed effect), delivery mode (fixed effect), child age at time of stool sample in months (repeated measurement), time between defecation as reported by caretaker and fieldworker’s retrieval of diaper in hours (repeated measurement), and maternal education (fixed effect). All models considered random effects by study participant for whom we had matched samples at 3, 6, 7, 9, 12 and 16 months of age. P-values were corrected for false discovery rate (FDR) using the Benjamini-Hochberg method to account for multiple hypothesis testing.

As sensitivity analyses, we evaluated the effect of household ownership of chickens, a possible source of ARG exposure,^28,29^ or weight-for-height z-scores, an indicator of child growth,^30^ on the association between total FPKM and antibiotic use. The effect estimates did not significantly change upon additional adjustments so we did not consider these in our final analyses (**Table S5**).

Next, we examined the effect of recent antibiotic use on the gut microbiome as measured by (1) richness; (2) Shannon diversity index; and (3) abundance of gut bacteria genera. Different taxonomic tools tend to yield discordant results at the species level;^31^ thus, we investigated effects at the genera level to balance granularity with reliability. Effects on richness and diversity were examined using separate adjusted linear mixed models. To examine effects on gut genera, we used linear mixed model-LinDA,^32^ a flexible statistical approach for correlated microbiome data with longitudinal measures. LinDA performs regression analysis on centered log2-ratio-transformed abundance data; identifies a bias term due to transformational and compositional effect; then uses the mode of the effect estimates (i.e., log2 fold-change) across different taxa to correct the effect estimates for the bias. All models controlled for the same covariates as described previously.

Lastly, we explored whether any observed effects of recent antibiotic use on ARG abundance may have been mediated by impacts on gut genera. Associations between the abundance of the ARGs that appeared to be impacted by antibiotic use (FDR<0.1) and the abundance of specific genera that were also impacted by the same antibiotic (FDR <0.1) were assessed using linear mixed models while adjusting for child age, time between defecation and diaper retrieval and study participant’s random effect.

All analyses were performed in R version 4.3.0 or above. Statistical significance was defined by α=0.05 and *p*-values are two-sided unless stated otherwise.

## Results

### Patterns of recent antibiotic use

Amoxicillin, cefalexin, sulfa-trimethoprim and azithromycin were the most used antibiotics (**Table 1**), although caretakers also reported the use of ampicillin, erythromycin, furazolidone, metronidazole, nifuroxazide, amikacin, cefaclor, cefradine, cefuroxime, and clarithromycin (**Table S1**). Sociodemographic-related differences in the number of antibiotic courses that children used in the 30 days prior to stool sampling have been summarized previously^23^ but briefly, it did not differ by age (**Table 1**), sex, delivery mode, maternal education or toilet type but differed by household water source (**Table S2**).

**Table 1.**
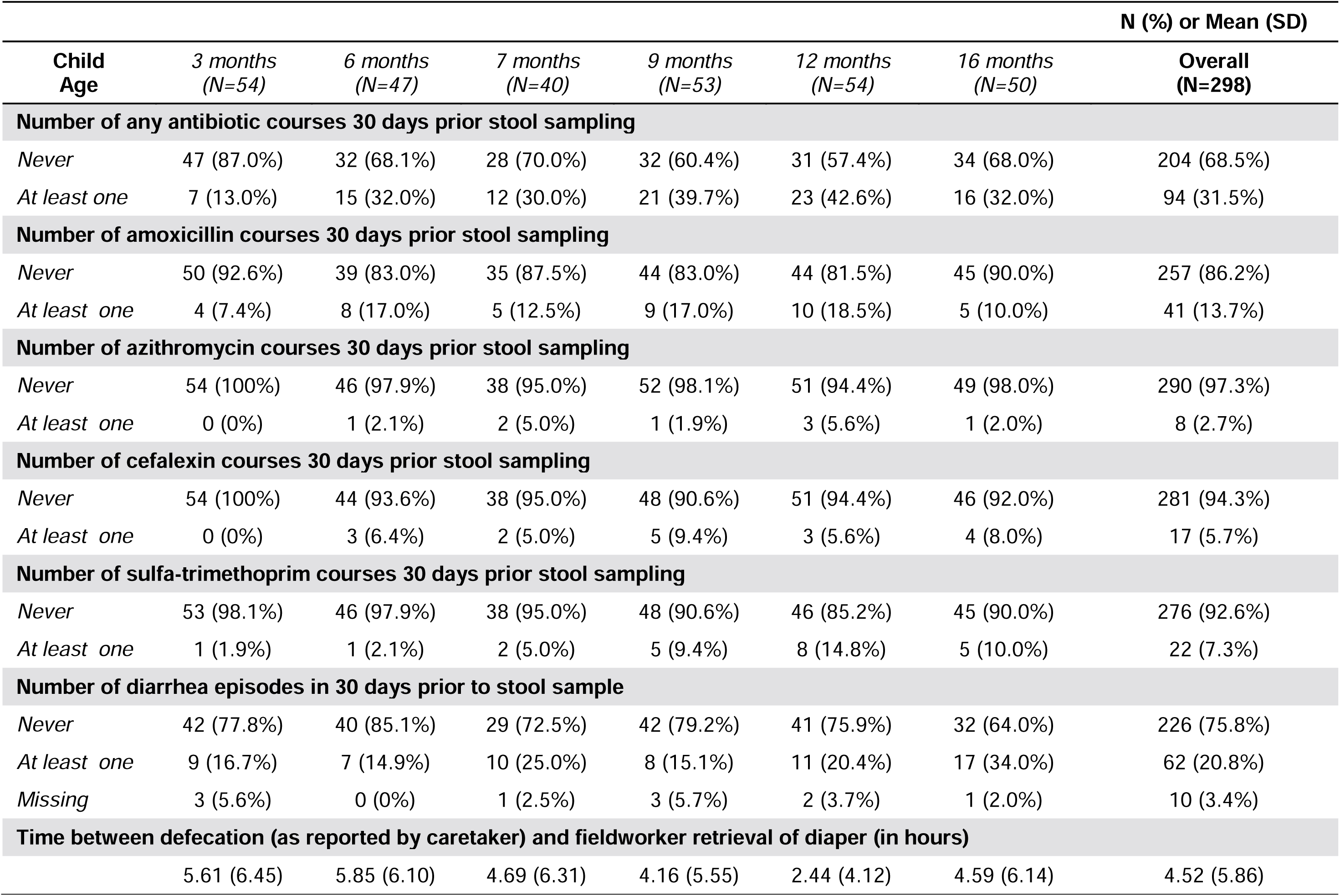
Patterns of recent antibiotic use and diarrhea among Peruvian children aged 3-16 months (N=54, repeated measures N=298) (2016-2019). *Abbreviations: SD, standard deviation*

|  | N (%) or Mean (SD) |  |  |  |  |  |  |
| --- | --- | --- | --- | --- | --- | --- | --- |
| **Child Age** | **3 months**<br>**(N=54)** | **6 months**<br>**(N=47)** | **7 months**<br>**(N=40)** | **9 months**<br>**(N=53)** | **12 months**<br>**(N=54)** | **16 months**<br>**(N=50)** | **Overall**<br>**(N=298)** |
| **Number of any antibiotic courses 30 days prior stool sampling** |  |  |  |  |  |  |  |
| *Never* | 47 (87.0%) | 32 (68.1%) | 28 (70.0%) | 32 (60.4%) | 31 (57.4%) | 34 (68.0%) | 204 (68.5%) |
| *At least one* | 7 (13.0%) | 15 (32.0%) | 12 (30.0%) | 21 (39.7%) | 23 (42.6%) | 16 (32.0%) | 94 (31.5%) |
| **Number of amoxicillin courses 30 days prior stool sampling** |  |  |  |  |  |  |  |
| *Never* | 50 (92.6%) | 39 (83.0%) | 35 (87.5%) | 44 (83.0%) | 44 (81.5%) | 45 (90.0%) | 257 (86.2%) |
| *At least one* | 4 (7.4%) | 8 (17.0%) | 5 (12.5%) | 9 (17.0%) | 10 (18.5%) | 5 (10.0%) | 41 (13.7%) |
| **Number of azithromycin courses 30 days prior stool sampling** |  |  |  |  |  |  |  |
| *Never* | 54 (100%) | 46 (97.9%) | 38 (95.0%) | 52 (98.1%) | 51 (94.4%) | 49 (98.0%) | 290 (97.3%) |
| *At least one* | 0 (0%) | 1 (2.1%) | 2 (5.0%) | 1 (1.9%) | 3 (5.6%) | 1 (2.0%) | 8 (2.7%) |
| **Number of cefalexin courses 30 days prior stool sampling** |  |  |  |  |  |  |  |
| *Never* | 54 (100%) | 44 (93.6%) | 38 (95.0%) | 48 (90.6%) | 51 (94.4%) | 46 (92.0%) | 281 (94.3%) |
| *At least one* | 0 (0%) | 3 (6.4%) | 2 (5.0%) | 5 (9.4%) | 3 (5.6%) | 4 (8.0%) | 17 (5.7%) |
| **Number of sulfa-trimethoprim courses 30 days prior stool sampling** |  |  |  |  |  |  |  |
| *Never* | 53 (98.1%) | 46 (97.9%) | 38 (95.0%) | 48 (90.6%) | 46 (85.2%) | 45 (90.0%) | 276 (92.6%) |
| *At least one* | 1 (1.9%) | 1 (2.1%) | 2 (5.0%) | 5 (9.4%) | 8 (14.8%) | 5 (10.0%) | 22 (7.3%) |
| **Number of diarrhea episodes in 30 days prior to stool sample** |  |  |  |  |  |  |  |
| *Never* | 42 (77.8%) | 40 (85.1%) | 29 (72.5%) | 42 (79.2%) | 41 (75.9%) | 32 (64.0%) | 226 (75.8%) |
| *At least one* | 9 (16.7%) | 7 (14.9%) | 10 (25.0%) | 8 (15.1%) | 11 (20.4%) | 17 (34.0%) | 62 (20.8%) |
| *Missing* | 3 (5.6%) | 0 (0%) | 1 (2.5%) | 3 (5.7%) | 2 (3.7%) | 1 (2.0%) | 10 (3.4%) |
| **Time between defecation (as reported by caretaker) and fieldworker retrieval of diaper (in hours)** |  |  |  |  |  |  |  |
|  | 5.61 (6.45) | 5.85 (6.10) | 4.69 (6.31) | 4.16 (5.55) | 2.44 (4.12) | 4.59 (6.14) | 4.52 (5.86) |

Nine children took more than one antibiotic course in the 30 days before a stool sample (**Figure S2**). Children either took the same antibiotic repeatedly (4/298 timepoints for 4/54 children) or combined antibiotics (10/298 timepoints for 9/54 children), most frequently sulfa-trimethoprim with erythromycin, amoxicillin, or azithromycin.

### Effect of recent antibiotic use on richness, diversity, and abundance of gut genera

Across all timepoints, *Bifidobacterium* (44.85%), *Blautia* (3.12%), *Bacteroides* (2.09%), and *Escherichia* (1.94%) were some of the most abundant genera on average.^23^ Species richness and Shannon diversity increased as children aged. After covariate adjustments, recent antibiotic use did not significantly affect species richness or diversity over the first 16 months of life (all p-values>0.05; **Table S3**).

The number of amoxicillin, azithromycin, cefalexin, and sulfa-trimethoprim courses recently used were not significantly associated with the abundance of any genera (FDR>0.05) but there were several notable trends (**Figure 1**). Azithromycin use was associated with decreased abundance of *Clostridium* (log2 fold-change=-0.71; FDR=0.18), *Roseburia* (log2 fold-change=-0.64; FDR=0.18), unclassified *Firmicutes* (log2 fold-change=-0.64; FDR=0.18) and *Dorea* (log2 fold-change=-0.47; FDR=0.18), but increased *Enterococcus* (log2 fold-change=0.76; FDR=0.18). Cefalexin use was associated with increased *Intestinibacter* abundance (log2 fold-change=0.44; FDR=0.05). Sulfa-trimethoprim was associated with decreased abundance of potential pathogens like *Klebsiella* (log2 fold-change=-0.59; FDR=0.14), *Citrobacter* (log2 fold-change=-0.36; FDR=0.17), and *Tyzzerella* (log2 fold-change=-0.50; FDR=0.12), and commensals such as *Eubacterium* (log2 fold-change=-0.82; FDR=0.07), *Parabacteroides* (log2 fold-change=-0.61; FDR=0.07); *Bifidobacterium* (log2 fold-change=-0.39, FDR=0.12), *Akkermansia* (log2 fold-change= –0.64, FDR=0.14); *Ruminococcus* (log2 fold-change=-0.55, FDR=0.12), and *Coprococcus* (log2 fold-change=-0.33, FDR=0.18).

**Figure 1.**
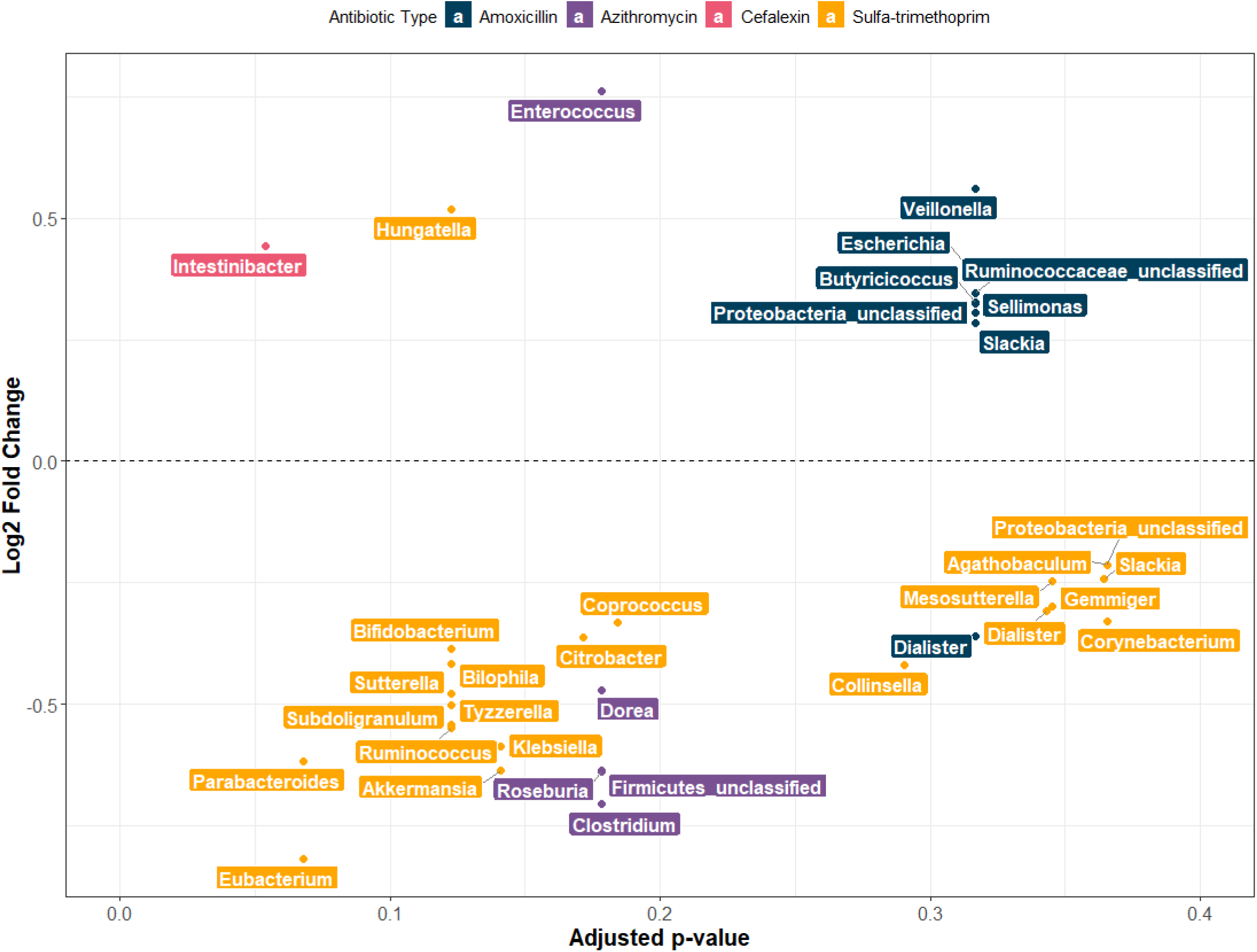
Differentially abundant bacterial genera following recent antibiotic use (past 30 days) among ruvian children aged 3-16 months (2016-2019). *Note: All models included repeated measures N=282 and adjusted for number of diarrhea episodes in 30 days prior to stool sample, child sex, delivery mode, child age at time of stool sample (in months), maternal education, time between defecation and diaper retrieval (in hours)*.

### Effect of recent antibiotic use on ARG abundance

On average, log10-transformed, normalized abundance of total ARGs in children’s stool ranged between 5.50 and 8.10 FPKM. We observed no sex-specific differences or statistically significant trends over the first 16 months of life (**Figure 2** and **S3**).

**Figure 2.**
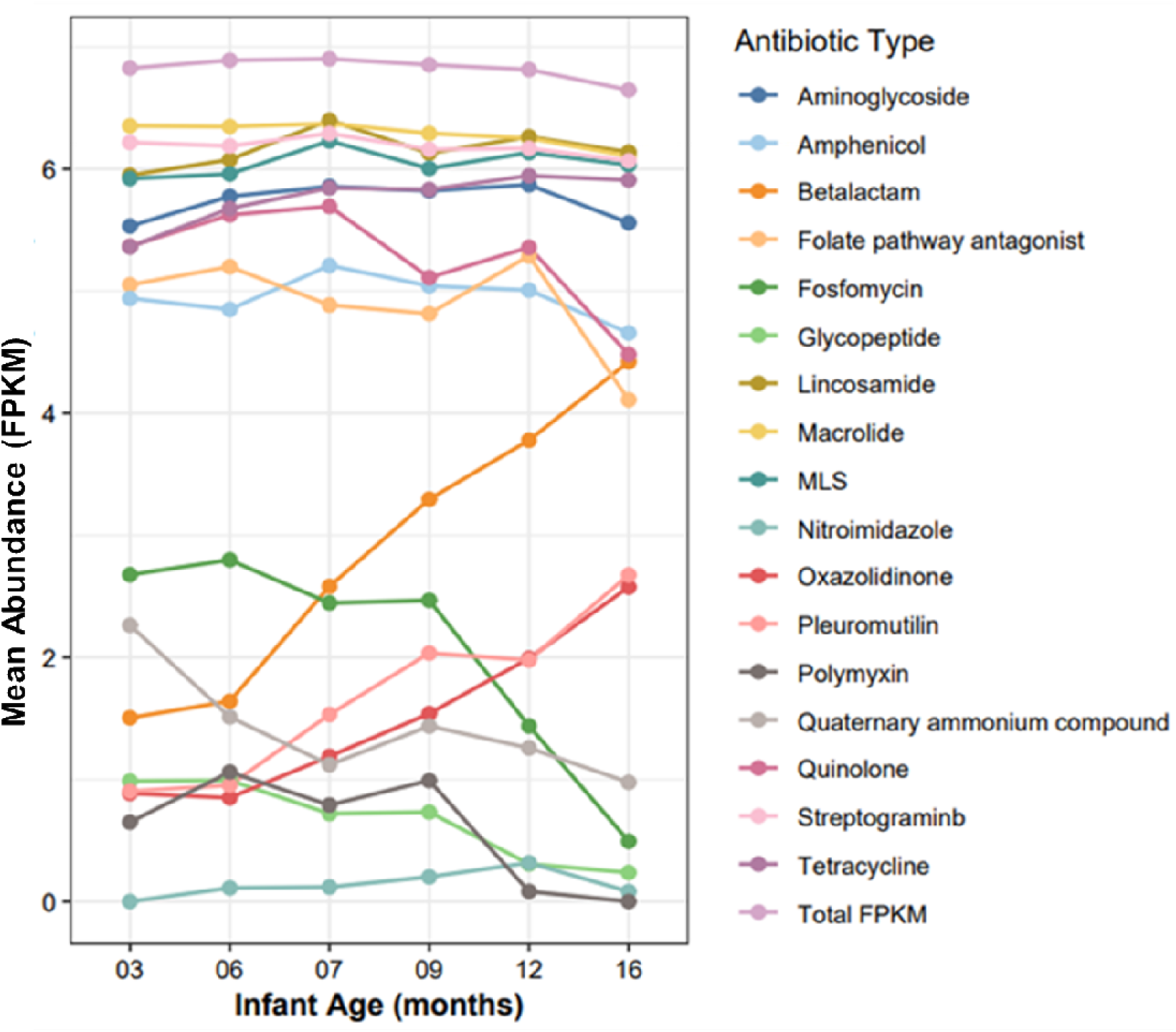
Age-stratified antibiotic resistance gene (ARG) abundance, overall and by class, among stool Samples from 54 Peruvian children aged 3-16 months (2016-2019). *Abbreviations: ARG, antibiotic resistance genes; FPKM, fragments per kilobase of transcript per million ents mapped*

All commonly used antibiotics except cephalexin significantly impacted some ARG abundances (**Figure 3**). Notably, ARGs with altered abundance did not typically confer resistance to the antibiotic that induced the observed effect. Increased amoxicillin use was significantly associated with increased load of *dfrA8* (conferring resistance to trimethoprim) (β=1.57; 95%CI=0.79,2.35). Azithromycin use decreased *aph(3’’)-Ib* (β=-2.64; 95%CI=-4.24,-1.04) abundance, but was associated with higher abundance of aminoglycoside resistance genes including *aadD* (β=1.83; 95%CI=0.66,3.00), *aadA2* (β=2.72; 95%CI=1.35,4.08), and *ant(9)-Ia* (β=2.83; 95%CI=1.35,4.31), as well as *dfrA12* (β=2.98; 95%CI=1.65,4.31) (trimethoprim resistance). Increased sulfa-trimethoprim use was linked with higher abundance of *lnu(B)* (β=1.32; 95%CI=0.69,1.95) (lincosamide resistance). Notably, but not significantly (FDR<0.10), amoxicillin was associated with higher abundance of *dfrA8* and *fosA6* (fosfomycin resistance), while azithromycin decreased abundance of *aph(6)-Id* (aminoglycoside resistance) but increased abundance of *erm(A)* (macrolide resistance).

**Figure 3.**
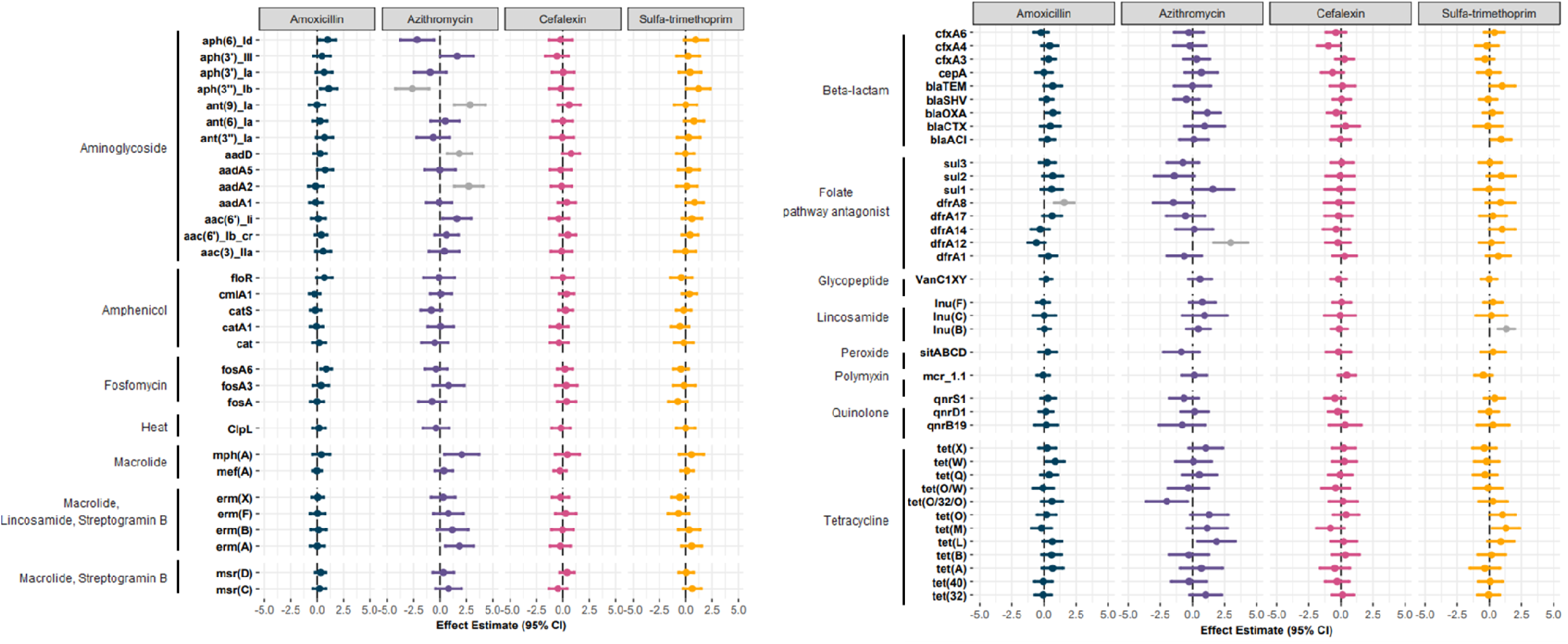
Associations between recent antibiotic use (past 30 days) antibiotic resistance gene (ARG) abundance in stool samples of 54 Peruvian children aged 3-16 months (2016-2019). *Note: All models included N=282 and adjusted for number of diarrhea episodes in 30 days prior to stool sample,* child *sex, delivery mode,* child *age at time of stool sample (in months), maternal education, time between defecation and diaper retrieval (in hours). ARG abundance was modeled as log_10_FPKM. Associations significant based on a false discovery rate of 0.05 are colored in grey*. *Abbreviations: ARGs, antibiotic resistance genes; FPKMs, fragments per kilobase of transcript per million fragments mapped*

Antibiotic use also exerted effects on ARG class (**Figure 3**). Amoxicillin was associated with significant increases in the abundance of folate pathways antagonist (β=0.76; 95%CI=0.14,1.38), aminoglycoside (β=0.43; 95%CI=0.07,0.80) and tetracycline ARGs (β=0.31; 95%CI=0.01,0.60). Cefalexin was associated with higher abundance of quaternary ammonium compounds ARGs (β=1.28; 95%CI=0.25,2.31), and notably, but non-significantly, lower abundance of β-lactam ARGs (β=-1.02; 95%CI=-2.05,0.01). Sulfa-trimethoprim use significantly decreased fosfomycin ARG abundance (β=-1.28; 95%CI=-2.48,-0.07). Only amoxicillin use increased total FPKM (β=0.15, 95%CI=0.01,0.29).

### Relation between abundance of ARGs and gut genera

Antibiotic use can drive increases in ARG abundance by either enriching ARGs among existing gut bacterial communities due to horizontal gene transfer or by increasing the abundance of bacteria that naturally or frequently harbor ARGs. To explore which mechanism could underlie the observed associations between antibiotic use and ARG abundances, we identified ARG and genus that were impacted by the same antibiotic’s use (using FDR<0.1). Only sulfa-trimethoprim use impacted both ARG and genus abundances, increasing *lnu(B)* (ARG) and decreasing *Eubacterium* and *Parabacteroides* (genera) abundance. However, the abundance of *lnu(B)* was not significantly associated with the abundances of *Eubacterium* (β=0.06; 95%CI=-0.32,0.43) or *Parabacteroides* (β=-0.11; 95%CI=-0.44,0.21) and so, it appeared unlikely that the increase in lnu(B) associated with sulfa-trimethoprim use was driven by specific gut genera. Because neither amoxicillin, azithromycin, nor cefalexin use affected any gut genera (based on FDR<0.1), their effects on specific ARGs (e.g., *dfrA8*, *dfrA12*, *fosA6, aph(3’’)-Ib*, *aadD, ant(9)-Ia*) appear to have been independent of effects on gut genera.

## Discussion

We examined the effects of four commonly used antibiotics (amoxicillin, azithromycin, cefalexin and sulfa-trimethoprim) on the gut microbiomes of Peruvian children raised in a peri-urban informal settlement of Lima. None of these antibiotics significantly altered the abundance of gut genera, species richness, nor species diversity over the first 16 months of life. However, amoxicillin, azithromycin, and sulfa-trimethoprim use significantly enriched ARGs, including those that do not confer resistance to the antibiotic that induced the effect. Given the lack of effects on gut genera or α-diversity, our analyses indicate that the effects of antibiotic use on the resistome might be independent of gut microbial changes among children in this setting. Overall, our findings underscore the dynamic and complex link between antibiotic use and development of the gut resistome in children and suggest that the increased use of common antibiotics may be key drivers of the development of the resistome among children in peri-urban Lima.

Prior literature suggests that increased consumption of antibiotics alters bacterial abundance in the gut as well as microbial richness^33,34^ and diversity^16,34^ and the abundance of its bacteria,^35^ but we did not observe this. This could be due to differences in the antibiotic types we studied, many of which are excreted in urine rather than feces (e.g., amoxicillin, cefalexin, sulfa-trimethoprim), and therefore may exert minimal effects on the gut microbiome. In addition, breastfeeding rates were exceptionally high in this study setting; 90% of children continued receiving breastmilk at 16 months of age. The effects of antibiotic use on the gut microbiome could have been less pronounced in our study because children’s gut microbiomes could ‘bounce-back’ or recover faster compared to children from other settings, who may have different breastfeeding exposures^36^ along with very frequent antibiotic usage^37^.

Azithromycin is the only antibiotic we examined that is primarily excreted in feces (half-life of 68 hours), and we consequentially noted substantial effects on children’s gut resistomes and non-significant but notable effects on the gut microbiome. Azithromycin is a semisynthetic macrolide used for treating community-acquired pneumonia, asthma, and periodontal infections in children.^49,50^ In addition to macrolide resistance genes, azithromycin significantly altered the abundance of ARGs conferring resistance to other classes, including aminoglycosides and folate pathways antagonists, possibly through the section of mobile genetic elements that co-encode resistance to azithromycin and other antibiotics. A study of children <5 years old in Burkina Faso found that 2 weeks after an oral azithromycin dose, the load of macrolide resistance determinants increased and gut diversity decreased; however, by 6 months, these effects were no longer distinguishable.^38^ The MORDOR trial in Niger, Malawi and Tanzania also showed that following azithromycin use by pre-schoolers, macrolide resistance increased.^39^ ^31,36^ Amoxicillin was the most commonly used antibiotic in this study. It did not significantly impact gut genera or α-diversity but significantly enriched the total ARG load in children’s guts.

Amoxicillin is a broad-spectrum penicillin derivative commonly prescribed for bacterial infections of the tonsils, lungs, ear, and urinary tract.^40^ It has a half-life of ∼1 hour and is primarily excreted through urine. Children treated with amoxicillin in a Niger trial had an enriched resistome, and enrichment of *Escherichia* but depletion of *Dorea* in the gut.^41^ Pre– and term-infants from the Netherlands with intravenous administration of amoxicillin/ceftazidime during the first 2 weeks of life had decreased *Escherichia-Shigella* abundance.^42^ *Bifidobacterium* are generally sensitive to amoxicillin^43–46^; however, we did not observe this, perhaps due to differences in dosing^47^ or high rates of breastfeeding in our population, which is a protective factor of the microbiome and may have helped maintain *Bifidobacterium* abundance. Inconsistencies with prior work could also be because we limited our analysis to the effects of amoxicillin use alone and not in combination with clavulanic acid, which exacerbates amoxicillin’s effects on the microbiome and possibly the resistome.^48^

Cefalexin is a β-lactam antibiotic most often prescribed for urinary tract infections.^49^ Cefalexin has a half-life of <2hrs and is largely unmetabolized before being excreted in urine. Here, cefalexin use marginally increased the abundance of *Intestinibacter*, a gut commensal suspected to play a role in glucose and lipid metabolism^50,51^, and significantly increased the abundance of quaternary ammonium compounds ARGs. Few studies have explored the consequences of early childhood exposure to cefalexin or cephalosporins on the resistome and gut microbiome. A study investigating the impact of postnatal oral ceflazin use on children’s fecal bacterial composition found that in the month post-treatment (similar to the time frame we examined), the microbiome did not significantly differ among infants who were treated versus who were not.^52^ Others have found that cephalosporin (e.g., cefprozil and cefpodoxime) use can alter microbiomes and select β-lactamase resistance genes^53–55^; however, these studies have been conducted among healthy adults and may not align with the effects among children as their gut microbiomes and resistomes are not yet stable.

Sulfa-trimethoprim or cotrimoxazole is used for treating bronchitis and diarrhea in children.^56^ Similar to the other antibiotics we analyzed, it is excreted in urine and has a half-life of 10hrs. In our study, its use depleted numerous gut commensals and potential pathogens (e.g., *Klebsiella*, *Citrobacter*), albeit not significantly. Prior studies suggest that cotrimoxazole can increase ARG abundance^57,58^ and deplete gut bacteria.^58,59^

Interestingly, we noted that some children used different antibiotic combinations in the 30 days before a stool sample. Simultaneous use of multiple antibiotics (polypharmacy) can occur in in-patient settings to delay the spread of antibiotic resistance or more effectively treat a resistant infection; however, the reasons for polypharmacy in the community setting are unclear. There may be unique effects of simultaneous broad-spectrum antibiotic use on microbiomes and resistomes which we were unable to assess due to low frequency of polypharmacy in this cohort.

Our study has many strengths. Leveraging weekly stool sampling and daily surveillance data, we were able to longitudinally analyze the effects of common antibiotics on the resistome and gut genera. Unlike most studies examining the effects of antibiotic use on child health, we expect that recall bias for antibiotic use was minimal, given that data were collected daily, and fieldworkers were able to confirm with medication packaging. By using LinDA^32^, we were able to correct our estimates for transformational and compositional bias. We expect our results to be minimally impacted by batch effects as all metagenomic sequencing was completed in two batches.

Our study has some limitations. First, our analyses do not indicate causality. The antibiotic-related effects on ARGs that we report may not necessarily translate to changes in expressed phenotypic resistance. Relatedly, because we did not perform functional genomics, we may have missed the effects of antibiotic use on novel ARGs. As with all epidemiological studies, our results are affected by residual confounding. For example, we were unable to account for diet-driven microbial changes after children began consuming solid foods. Because children in this study were mostly breastfed and breastmilk samples were not collected, we were unable to study effect modification by breastmilk intake or composition, which might be a source of bacteria^60–62^ and ARGs^63^. Additionally, we were unable to examine how prenatal exposures^7,13^ or the maternal microbiome^45,64–66^ may influence children’s gut microbiomes. Lastly, we selected children based on ESBL-E gut colonization patterns and thus our results may not be generalizable to all children in peri-urban Lima.

Nonetheless, to our knowledge, our study is the first look at antibiotic-specific effects on the gut microbiome and resistome among Peruvian children. Future studies should examine how polypharmacy, chemical exposures (e.g., quaternary ammonium compounds), and maternal exposures (e.g., breastmilk and stress) modify the associations between antibiotic use, and the gut microbiome and resistome.

## Acknowledgments

We are grateful to the families, especially the caretakers and their children, who were willing to participate in this cohort. We also are grateful to the study’s fieldworkers and team for their dedication, collaboration, and support.

## Funding

This work was supported by NIH R01AI108695-01A, KL2TR002545, U19AI110818 and 5T32ES012870. MLN was supported by Emory University and the MP3 Initiative. The content is solely the responsibility of the authors and does not necessarily represent the official views of Emory University or the MP3 Initiative.

## Transparency declarations

The authors have no conflicts of interest.

## Data availability

Children’s fecal metagenomes are available in NCBI’s Sequence Read Archive (https://www.ncbi.nlm.nih.gov/sra) under BioProject number PRJNA1138246.

## Notes

### Competing Interest Statement

The authors have declared no competing interest.

### Author Declarations

Infants' caretakers provided written informed consent for participation in the parent cohort and the use of collected specimens for subsequent research. The Institutional Review Boards (IRBs) of the Universidad Peruana Cayetano Heredia (UPCH), Johns Hopkins University and Asociacion Benefica PRISMA approved the parent study. Analyses for this sub-study were approved by the IRBs of UPCH (no. 201592), PRISMA, and Tufts University.

