## Supplemental File for "Effects of commonly used antibiotics on children’s developing gut microbiomes and resistomes in peri-urban Lima, Peru"

**Table S1. Recent antibiotic use (past 30 days) among 54 Peruvian children aged 3-16 months (2016-2019).**

|  | **Overall**  **(N=298)** |
| --- | --- |
| **Number of cefradine courses 30 days prior stool sampling** | |
| Never | 297 (99.7%) |
| At least once | 1 (0.3%) |
| **Number of furazolidone courses 30 days prior stool sampling** | |
| Never | 292 (98.0%) |
| At least once | 6 (2.0%) |
| **Number of amoxicillin con ac clavulanic courses 30 days prior stool sampling** | |
| Never | 285 (95.6%) |
| At least once | 13 (4.3%) |
| **Number of nifuroxazide courses 30 days prior stool sampling** | |
| Never | 298 (100%) |
| **Number of erythromycin courses 30 days prior stool sampling** | |
| Never | 295 (99.0%) |
| At least once | 3 (1.0%) |
| **Number of metronidazole courses 30 days prior stool sampling** | |
| Never | 298 (100%) |
| **Number of cefaclor courses 30 days prior stool sampling** | |
| Never | 297 (99.7%) |
| At least once | 1 (0.3%) |
| **Number of ampicillin courses 30 days prior stool sampling** | |
| Never | 296 (99.3%) |
| At least once | 2 (0.7%) |
| **Number of amikacin courses 30 days prior stool sampling** | |
| Never | 298 (100%) |
| **Number of clarithromycin courses 30 days prior stool sampling** | |
| Never | 297 (99.7%) |
| At least once | 1 (0.3%) |
| **Number of dicloxacillin courses 30 days prior stool sampling** | |
| Never | 298 (100%) |
| **Number of cefuroxime courses 30 days prior stool sampling** | |
| Never | 298 (100%) |

**Table S2. Summary of recent amoxicillin, azithromycin, cefalexin, and sulfa-trimethoprim use (past 30 days) among 54 Peruvian children aged 3-16 months (2016-2019), stratified by child sex, maternal education level, delivery mode, toilet type and water source.**

*Note: *Delivery mode was not reported for one child*

|  | Child Sex | | Maternal Education | | Delivery Mode* | | Toilet Type | | Household Water Source | | |
| --- | --- | --- | --- | --- | --- | --- | --- | --- | --- | --- | --- |
|  | Female | Male | At least high school | Less than high school | C-section | Vaginal birth | Pit Latrine | Indoor flush toilet | Public shared tap | 24hr indoor connection | Other |
|  | N=164 | N=134 | N=191 | N=107 | N=78 | N=214 | N=66 | N=232 | N=67 | N=216 | N=15 |
| Number of any antibiotic courses 30 days prior stool sampling | | | | | | | | | | | |
| Never | 113  (68.9%) | 91 (67.9%) | 127  (66.5%) | 77  (72.0%) | 56  (71.8%) | 143  (66.8%) | 52 (78.8%) | 152 (65.5%) | 55 (82.1%) | 141 (65.3%) | 8  (53.3%) |
| At least once | 51  (31.1%) | 43 (32.1%) | 64  (33.5%) | 30  (28.0%) | 22  (28.2%) | 71  (33.2%) | 14  (21.2%) | 80  (34.4%) | 12  (17.9%) | 75  (34.8%) | 7  (46.6%) |
| Number of amoxicillin courses 30 days prior stool sampling | | | | | | | | | | | |
| Never | 142  (86.6%) | 115 (85.8%) | 169  (88.5%) | 88  (82.2%) | 68  (87.2%) | 184  (86.0%) | 57 (86.4%) | 200 (86.2%) | 58 (86.6%) | 186 (86.1%) | 13 (86.7%) |
| At least once | 22  (13.4%) | 19 (14.2%) | 22  (11.5%) | 19  (17.8%) | 10  (12.8%) | 30  (14.0%) | 9 (13.6%) | 32  (13.8%) | 9 (13.4%) | 30  (13.9%) | 2  (13.3%) |
| Number of azithromycin courses 30 days prior stool sampling | | | | | | | | | | | |
| Never | 159 (97.0%) | 131 (97.8%) | 186  (97.4%) | 104  (97.2%) | 75  (96.2%) | 209  (97.7%) | 66 (100%) | 224 (96.6%) | 67 (100%) | 210 (97.2%) | 13 (86.7%) |
| At least once | 5  (3.0%) | 3  (2.2%) | 5  (2.6%) | 3  (2.8%) | 3  (3.8%) | 5  (2.3%) | 0  (0%) | 8  (3.4%) | 0  (0%) | 6  (2.8%) | 2  (13.3%) |
| Number of cefalexin courses 30 days prior stool sampling | | | | | | | | | | | |
| Never | 156 (95.1%) | 125 (93.3%) | 177  (92.7%) | 104  (97.2%) | 73  (93.6%) | 202  (94.4%) | 62 (93.9%) | 219 (94.4%) | 63  (94.0%) | 203 (94.0%) | 15  (100%) |
| At least once | 8  (4.9%) | 9  (6.7%) | 14  (7.3%) | 3  (2.8%) | 5  (6.4%) | 12  (5.6%) | 4  (6.1%) | 13  (5.6%) | 4  (6.0%) | 13  (6.0%) | 0  (0%) |
| Number of sulfa-trimethoprim courses 30 days prior stool sampling | | | | | | | | | | | |
| Never | 153 (93.3%) | 123 (91.8%) | 176  (92.1%) | 100  (93.5%) | 69  (88.5%) | 201  (93.9%) | 64 (97.0%) | 212 (91.4%) | 65 (97.0%) | 198 (91.7%) | 13  (86.7%) |
| At least once | 11  (6.7%) | 11  (8.2%) | 15  (7.9%) | 7  (6.5%) | 9  (11.5%) | 13  (6.1%) | 2  (3.0%) | 20  (8.6%) | 2  (3.0%) | 18  (8.3%) | 2  (13.4%) |

**Table S3. Unadjusted and adjusted estimates of the associations of recent amoxicillin, azithromycin, cefalexin, or sulfa-trimethoprim used (past 30 days) with gut species richness and Shannon diversity among 54 Peruvian children aged 3-16 months (2016-2019).**

*Note: Unadjusted models accounted for repeat measures from each child. Adjusted models additionally included number of diarrhea episodes in 30 days prior to stool sample, child sex, delivery mode, child age at time of stool sample (in months), maternal education, time between defecation and diaper retrieval (in hours).*

*Abbreviations:* *β, effect estimates; CI, confidence intervals*

|  | *N* | Log10 transformed Species Richness | Shannon Diversity Index |
| --- | --- | --- | --- |
|  |  | **β (95%CI)** | **β (95%CI)** |
| Unadjusted Models | | | |
| Number of amoxicillin courses 30 days prior to stool sampling | 282 | 0.02  (-0.06, 0.09) | 0.15  (-0.11, 0.41) |
| Number of azithromycin courses 30 days prior to stool sampling | 282 | -0.05  (-0.20, 0.11) | -0.08  (-0.60, 0.44) |
| Number of cefalexin courses 30 days prior to stool sampling | 282 | 0.03  (-0.08, 0.13) | 0.20  (-0.15, 0.56) |
| Number of sulfa-trimethoprim courses 30 days prior to stool sampling | 282 | 0.10  (-0.00, 0.19) | 0.40  (0.07, 0.73) |
| Adjusted Models | | | |
| Number of amoxicillin courses 30 days prior to stool sampling | 282 | 0.02  (-0.04, 0.07) | 0.13  (-0.05, 0.31) |
| Number of azithromycin courses 30 days prior to stool sampling | 282 | -0.09  (-0.19, 0.02) | -0.19  (-0.55, 0.17) |
| Number of cefalexin courses 30 days prior to stool sampling | 282 | -0.02  (-0.09, 0.05) | 0.03  (-0.22, 0.28) |
| Number of sulfa-trimethoprim courses 30 days prior to stool sampling | 282 | -0.01  (-0.08, 0.06) | 0.00  (-0.25, 0.25) |

**Table *S*4. Adjusted differential abundance analysis of bacterial genera and recent anitbiotic use (past 30 days) among 54 Peruvian children aged 3-16 months (2016-2019).**

*Note: Models with false discovery rate or adjusted p-value<0.4 are shown. All models included repeated N=282. All models adjusted for number of diarrhea episodes in 30 days prior to stool sample, child sex, delivery mode, child age at time of stool sample (in months), maternal education, time between defecation and diaper retrieval (in hours).*

| Antibiotic  Used | Gut Microbiome  Genera | Differential Abundance Analysis | | |
| --- | --- | --- | --- | --- |
|  |  | **log2 Fold Change** | **Statistic** | **Adjusted**  **p-value** |
| Number of amoxicillin courses 30 days prior stool sampling | Veillonella | 0.56 | 2.28 | 0.32 |
|  | Escherichia | 0.38 | 2.42 | 0.32 |
|  | Butyricicoccus | 0.33 | 2.11 | 0.32 |
|  | unclassified Ruminococcaceae | 0.35 | 2.18 | 0.32 |
|  | Slackia | 0.29 | 2.09 | 0.32 |
|  | Sellimonas | 0.32 | 2.12 | 0.32 |
|  | unclassified Proteobacteria | 0.31 | 2.42 | 0.32 |
|  | Dialister | -0.36 | -2.21 | 0.32 |
| Number of azithromycin courses 30 days prior stool sampling | Clostridium | -0.71 | -2.50 | 0.18 |
|  | unclassified Firmicutes | -0.64 | -2.52 | 0.18 |
|  | Dorea | -0.47 | -2.62 | 0.18 |
|  | Enterococcus | 0.76 | 2.59 | 0.18 |
|  | Roseburia | -0.64 | -2.64 | 0.18 |
| Number of cefalexin courses 30 days prior stool sampling | Intestinibacter | 0.44 | 3.40 | 0.05 |
| Number of sulfa-trimethoprim courses 30 days prior stool sampling | Bifidobacterium | -0.39 | -2.47 | 0.12 |
|  | Corynebacterium | -0.33 | -1.59 | 0.37 |
|  | Collinsella | -0.42 | -1.89 | 0.29 |
|  | Akkermansia | -0.64 | -2.29 | 0.14 |
|  | Klebsiella | -0.59 | -2.32 | 0.14 |
|  | Tyzzerella | -0.50 | -2.69 | 0.12 |
|  | Hungatella | 0.52 | 2.51 | 0.12 |
|  | Agathobaculum | -0.21 | -1.59 | 0.37 |
|  | Parabacteroides | -0.62 | -3.22 | 0.07 |
|  | Eubacterium | -0.82 | -3.12 | 0.07 |
|  | Ruminococcus | -0.55 | -2.66 | 0.12 |
|  | Sutterella | -0.48 | -2.42 | 0.12 |
|  | Coprococcus | -0.33 | -2.12 | 0.18 |
|  | Subdoligranulum | -0.54 | -2.43 | 0.12 |
|  | Gemmiger | -0.30 | -1.72 | 0.35 |
|  | Slackia | -0.24 | -1.67 | 0.36 |
|  | Bilophila | -0.42 | -2.51 | 0.12 |
|  | Citrobacter | -0.36 | -2.18 | 0.17 |
|  | unclassified Proteobacteria | -0.21 | -1.60 | 0.37 |
|  | Mesosutterella | -0.25 | -1.74 | 0.35 |
|  | Dialister | -0.31 | -1.78 | 0.34 |

**Table S5. Sensitivity analysis examining if inclusion of household ownership of chicken or weight-for-height z-scores changes the effect estimate for the association between recent antibiotic use (past 30 days) and abundance of total FPKM for the 54 Peruvian children aged 3-16 months (2016-2019).**

*Note: All models adjusted for number of diarrhea episodes in the 30 days prior to stool sample, child sex, delivery mode, child age at time of stool sample (in months), maternal education, time between defecation and diaper retrieval (in hours).*

*Abbreviations: ARG, antibiotic resistance genes; CI, confidence intervals; FPKM, fragments per kilobase of transcript per million fragments mapped*

|  | *N* | Adjusted Model for total FPKM | Adjusted model for total FPKM with household ownership of chickens | Adjusted Model for total FPKM with weight for height z-scores |
| --- | --- | --- | --- | --- |
|  |  | **β (95%CI)** | **β (95%CI)** | **β (95%CI)** |
| Number of amoxicillin courses 30 days prior to stool sampling | 282 | 0.15  (0.01, 0.29) | 0.15  (0.01, 0.29) | 0.15  (0.01, 0.29) |
| Number of azithromycin courses 30 days prior to stool sampling | 282 | 0.11  (-0.16, 0.39) | 0.11  (-0.16, 0.39) | 0.11  (-0.16, 0.39) |
| Number of cefalexin courses 30 days prior to stool sampling | 282 | -0.01  (-0.20, 0.18) | -0.01  (-0.20, 0.18) | -0.01  (-0.20, 0.18) |
| Number of sulfa-trimethoprim courses 30 days prior to stool sampling | 282 | 0.10  (-0.09, 0.29) | 0.10  (-0.09, 0.29) | 0.10  (-0.10, 0.29) |

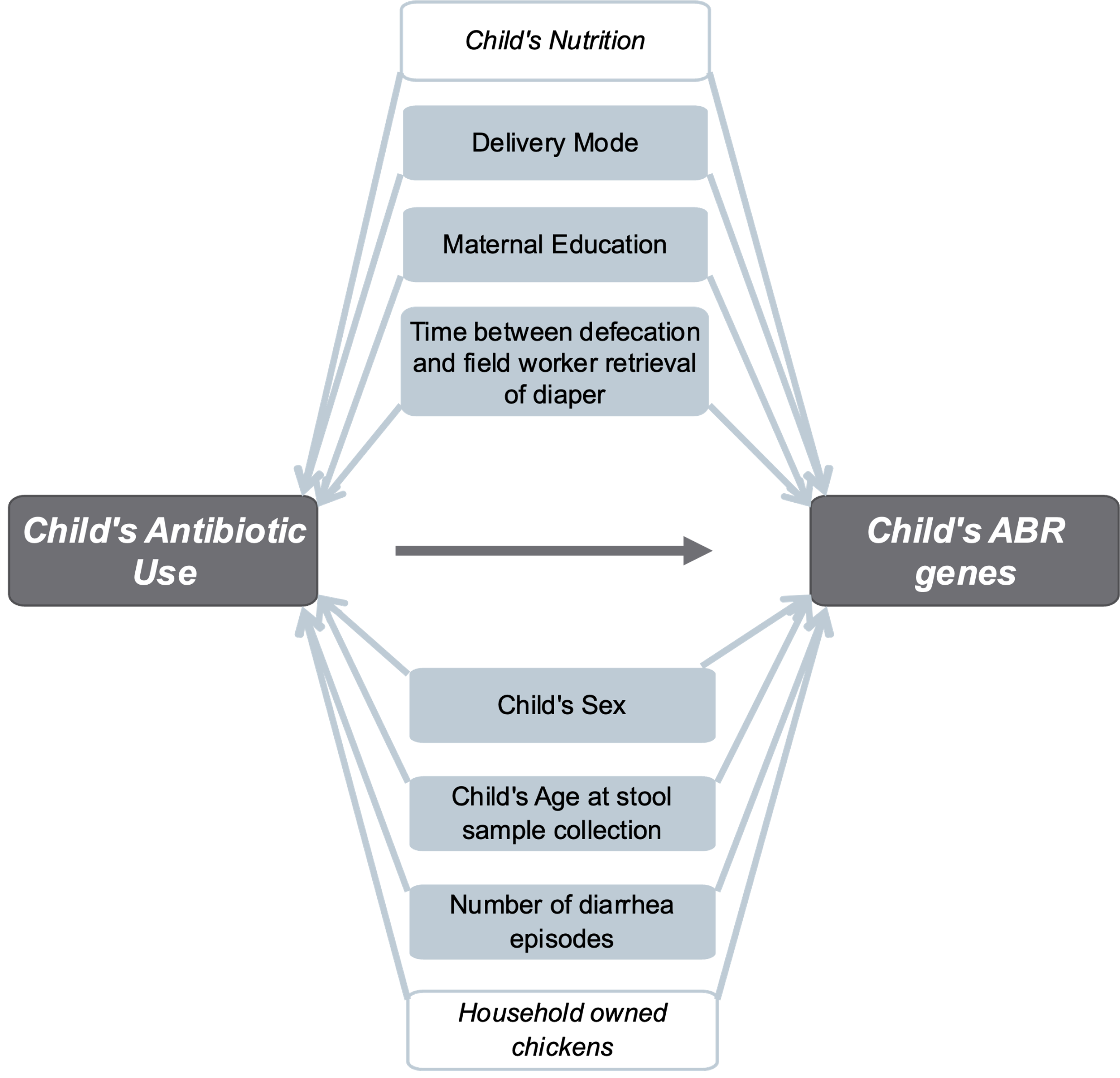

**Figure S1. Directed acyclic graph for the association between recent antibiotic use and load of antibiotic resistance genes in gut among Peruvian children aged 3-16 months (2016-2019).**

*Caption: Grey boxes depict the main exposure and outcome relationship. Variables in blue boxes were considered as important confounders. White boxes depict the variables that were considered as potential confounders and for whom the effect was test using sensitivity analysis.*

*Abbreviations: ABR, antibiotic resistance*

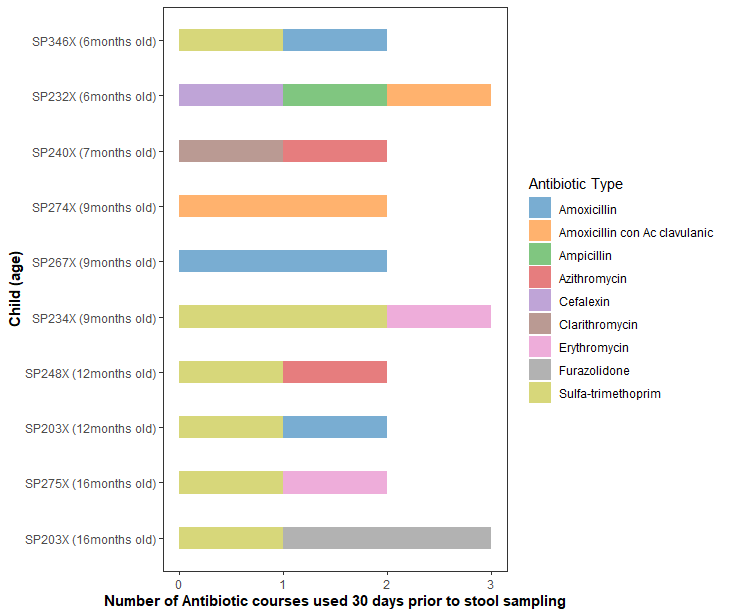

**Figure S2. Combinations of recent antibiotic use (past 30 days) among Peruvian children aged 3-16 months (2016-2019).**

*Note: Child SP203X appears twice since caretakers reported the child’s exposure to multiple antibiotics at 12 and 16 months of age.*

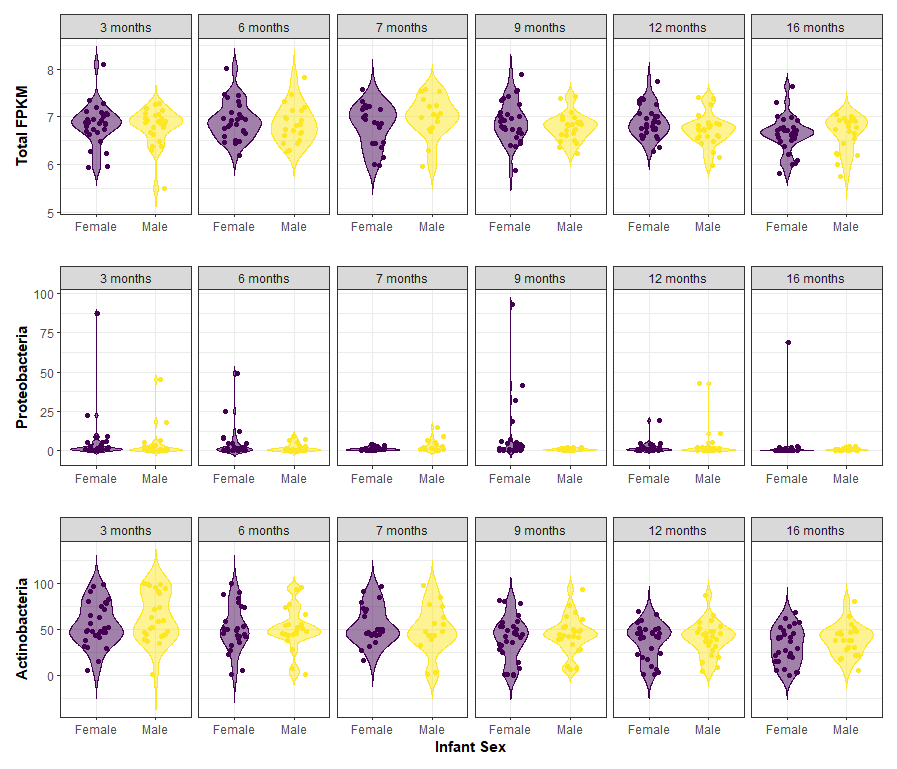

**Figure S3. Distribution of log10-transformed, normalized abundance of antibiotic resistance genes (measured by total FPKM) in the stool of 54 Peruvian children aged 3-16 months, stratified by child sex (2016-2019).**

*Note: To normalize for total bacteria in each sample, FPKM were calculated using the total number of genome equivalent in each sample, as estimated using Microbe Census.*

*Abbreviations: FPKM, fragments per kilobase per million mapped reads.*

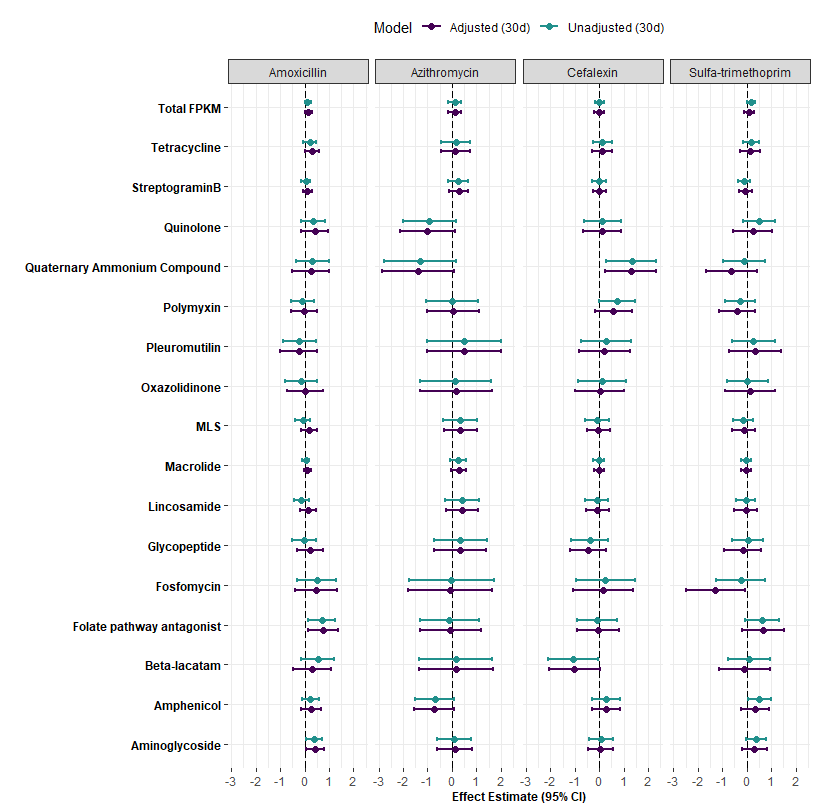

**Figure S4. Unadjusted and adjusted estimates of the association between recent amoxicillin, azithromycin, cefalexin, or sulfa-trimethoprim use (past 30 days) and abundance of ARGs (measured by FPKM), overall and by class among 54 Peruvian children aged 3-16 months (2016-2019).**

*Note: Unadjusted models accounted for repeat measures from each child. Adjusted models additionally included number of diarrhea episodes in 30 days prior to stool sample, child sex, delivery mode, child age at time of stool sample (in months), maternal education, time between defecation and diaper retrieval (in hours).*

*Abbreviations: ARGs, antibiotic resistance genes; FPKM, fragments per kilobase per million mapped reads*
